# Early Experience with COVD-19 Patients at Academic Hospital in Southwestern United States

**DOI:** 10.1101/2020.05.15.20094284

**Authors:** Rahul Shekhar, Shubhra Upadhyay, Abubaker Sheikh, Jeanette Atencio, Devika Kapuria

**Author notes:** Corresponding Author: Rahul Shekhar, MBBS, MD, 2211 Lomas BLVD, Department of internal medicine, University of New Mexico Health Science Center, Albuquerque, NM 87106. Corresponding author institutional. Alternative proof reader Devika Kapuria, MD. **Site of study:** This study was performed at University of New Mexico Health Science Center. **Author Contributions:** Drs Shekhar and Upadhyay had full access to all the data in the study and take responsibility for the integrity of the data and the accuracy of the data analysis. Both collected data. **Funding:** This study is not funded. **Study approval:** Study approval was obtained from the University of New Mexico Health Science Center Institutional Review Board and waiver of consent was given.

## Abstract

**Objective:** To describe the experience with COVID-19 patients hospitalized at University of New Mexico (UNM)

**Methods:** First 50 adults admitted at the UNM between 1/19 to 4/24 2020 were included in this retrospective study

**Results:** median age was 55 (20-85), 54% Female. Obesity (51%), diabetes (36%) and hypertension (34%) were most common comorbidities. Mean symptoms duration before admission 7days. Most common symptoms, subjective fevers (95.2%), cough (93%) and shortness of breath (87%). At triage 80% patients required oxygen, 20% required intubation. Differential 46% had elevated neutrophil counts and 48% had low lymphocytes counts. Median D dimer, Ferritin, CRP, LDH all elevated at presentation. 10% of patients had a negative chest x ray. 19.3% patients have coinfection with another respiratory pathogen. 68% patient required ICU level. 34% were directly admitted to ICU and 82.4% required invasive mechanical ventilation. Patients spent a median of 2 days on the floor prior to ICU transfer, median length of stay in the ICU-7 days. Patients with diabetes, and higher lactate dehydrogenase on admission were more likely to require ICU level of care. Of 34 patients in the ICU 13 died - overall mortality of 30.2%, Majority has AKI and required CRRT or HD. The median length of stay 7(Range 1-31days). 55% discharged home, 11% to rehabilitation facilities. 53.3% required oxygen on discharge.

**Conclusion:** This study provides early experience in treating patient admitted to Academic Hospital in state of New Mexico.

## Introduction

The first patient with coronavirus in the world was hospitalized on December 12. Since its initial emergence it has spread rapidly and as of May 4^th^ there have been three million cases and 247630 deaths [^1^]. The first case of coronavirus in the US was hospitalized on the 19th of January in Washington, after a trip to Wuhan, China[^2^], since then more than one million cases are reported in united states with 67682 deaths^1^

There is limited information available to describe complaints, natural history and clinical outcomes of hospitalized US patients and aim of this article is to present the early experience in treatment and management of COVID-19 patients in a tertiary care teaching hospital in southwestern United States to help educate health care professionals.

## Methods

We performed a retrospective and prospective cohort study of first 50 adults admitted at the University of New Mexico (UNM) Health Science center, the only tertiary care teaching hospital in the state of New Mexico. Study approval was obtained from the University of New Mexico Health Science Center Institutional Review Board and waiver of consent was given. A Chart review was performed for first 50 patients admitted to UNM between Jan 19^th^ to April 24^th^ 2020 who met the WHO case definition and had a confirmed positive result on polymerase chain reaction testing of a nasopharyngeal sample. Clinical outcomes were monitored until April 24^th^. Data was collected by 2 independent physicians (RS, SU) by extensive electronic medical record review. Data collected included patient demographics, comorbidities reported in history of presenting illness, travel history and family exposure history, social history, presenting vital signs, baseline laboratory and imaging results, inpatient medications, treatments, clinical course and complications as well as and outcomes. Clinical outcomes available for those in hospital at the study cut-off are presented, including invasive mechanical ventilation, intensive care unit (ICU) care, Kidney replacement therapy and length of stay in hospital. Outcomes such as discharge disposition and readmission were not available for patients in the hospital at study cut-off. Data was analyzed using STATA 16.1. Descriptive analysis was performed on all variables. Chi-squared analysis was performed comparing patients that were admitted to the floor vs. patients that were admitted to the ICU. Kaplan Meier survival graph was produced with endpoint being death comparing floor patient vs. ICU patients.

## Results

### Patient characteristics

At study censure, 43/50(86%) patients were discharged. Median age of patients was 55.5 years (range 20-85 years). Out of these 23(46%) were male and 27(54%) were female. 29/50(58%) were American Indian and 14/50 (28%) were white. (Table 1-11). 42/50(84%) had at least one comorbidity. Obesity was the most common comorbidity in 20/39 (51%), followed by diabetes in 18/50 (36%) and hypertension 17/50(34%).

Most common symptoms on presentation included subjective fevers 40/42 (95.2%), cough 43/46 (93%) 43/46 and shortness of breath 40/46(87%). Other symptoms reported were headache, malaise, myalgia, chest pain and gastrointestinal symptoms including nausea, vomiting, diarrhea and abdominal pain (table 1-2). Mean onset of symptoms duration before admission 7.39 days (range 1-21days). 28/45(62.2%) patients sought outpatient medical care (PCP/ER/urgent care) before admission. While 19/50(38%) patients were transferred to our hospital from outside hospital (OSH) for a higher level of care, only 2/50 (4%) patients were admitted from skilled nursing facilities (SNF).

**Table I:** Baseline Characteristics.

| Table I : Baseline Characteristics |  |
| --- | --- |
| Demographics | (n/N) |
| Number of patients | 50 |
| Age (median) [range] | (55.5) [20-85] |
| Sex |  |
| Male (%) | 23/50 (46) |
| Female (%) | 27/50 (54) |
| Ethnicity |  |
| Hispanic (%) | 14/50 (28) |
| Not-Hispanic (%) | 34/50 (68) |
| Race |  |
| White/Anglo | 14/50 (28) |
| American Indian/Alaska Native | 29/50 (58) |
| Black/African American | 1/50 (2) |
| Hawaiian Native or Pacific Islander | 1/50 (2) |
| Others | 1/50 (2) |
| Comorbidities | 42/50 (84) |
| Cardiovascular disease |  |
| Hypertension (%) | 17/50 (34) |
| CAD (%) | 3/50 (6) |
| CHF (%) | 2/50 (4) |
| CVA (%) | 2/50 (4) |
| Respiratory disease |  |
| Asthma (%) | 6/50 (12) |
| OSA (%) | 2/50 (4) |
| Liver disease |  |
| Non Alcoholic Liver disease | 2/50 (4) |
| Active alcohol user | 6/50 (12) |
| Alcoholic cirrhosis (%) | 2/50 (4) |
| Obesity | 20/39 (51) |
| BMI (median) [IQR] | (30.4) [ 25.7 - 35.1] |
| Class 1 (%) | 10/39 (26) |
| Class 2 (%) | 4/39 (10) |
| Class 3 (%) | 6/39 (15) |
| Diabetes (%) | 18/50 (36) |
| Smoking (%) | 24/50 (48) |
| Cancer history (%) | 4/50 (8) |

Legends: Coronary artery disease (CAD) , Congestive heart failure ( CHF ) , Cerebrovascular accident (CVA), obstructive sleep apnea (OSA), basal metabolic rate ( BMI )

### Clinical variables

Though 40/42 (95.2%) people complained of subjective fever only 12/50 (24%) were actually febrile (defined as temp > 38 degree Celsius) on triage vitals. Surprisingly 23/50 (46%) of patients did not have a single febrile episode throughout the hospitalization. 28/50 (56%) had a respiratory rate > 20 breaths per minute and 33/50 (66 %) had a heart rate more than 90 beats per minute on triage. 40/50 (80%) patients has oxygen saturations (Sats) less than 88% on room air (Ra) and required supplemental oxygen. 28/50 (56%) were initially treated with oxygen with nasal cannula, 4/50 (8%) required high flow nasal cannula, 1/50(2%) required bipap and 10/50(20%) were intubated. 34/50(68%) patients required ICU level care of care at some point during their hospitalization.

#### Laboratory and imaging characteristics

(Table II)

**Table II:** Admission symptoms, vitals and labs.

| **Table II : Admission symptoms, vitals and labs** |  |  |  |
| --- | --- | --- | --- |
|  | **All Patients** | **Floor** | **ICU** |
| Admission vitals | (n/N) | (n/N) | (n/N) |
| Systolic BP (Median) [Range] | (120) [60-172] | (133) [92 - 172] | (119) [ 60-170] |
| Diastolic BP (median) [Range] | (72.5) [37-110] | (83.8) [62-98] | (68.5) [37-110] |
| O2 sats |  |  |  |
| <88% (%) | 40/50 (80) | 10/16 (62) | 30/34 (88) |
| Febrile on admission (%) | 27/50 (54) | 7/16 (43) | 20/34 (59) |
| Median Temperature | 37.2 | 36.85 | 37.35 |
| Need supplemental oxygen |  |  |  |
| On Room Air (%) | 7/50 (14) | 5/16 (31) | 2/34 (6) |
| Nasal Canula (%) | 28/50 (56) | 11/16 (69) | 17 (34) 50 |
| High flow / Nonrebreather (%) | 4/50 (4) | 0/16 | 4/34 (12) |
| BIPAP (%) | 1/50 (2) | 0/16 | 1/34 (3) |
| Intubated (%) | 10/50 (2) | 0/16 | 10/34 (29) |
| Symptoms |  |  |  |
| Subjective fevers (%) | 40/42(95) | 14/14(100) | 26/29(93) |
| Cough (%) | 43/46 (93) | 14/15 (93) | 29/31 (93) |
| Shortness of Breath (%) | 40/46(87) | 13/15(87) | 27/32(87) |
| Chest pain (%) | 11/27 (41) | 5/13 (38) | 6/14 (43) |
| Nausea (%) | 10/28 (36) | 5/11 (45) | 5/17 (29) |
| Diarrhea (%) | 9/28 (32) | 4/12 (33) | 5/16 (31) |
| Vomiting (%) | 5/27 (18) | 2/11 (18) | 3/16 (19) |
| Sore throat (%) | 3/20 (15) | 1/8 (13) | 2/12 (17) |
| Abdominal pain (%) | 3/23 (13) | 1/10 (10) | 2/13 (15) |
| Runny Nose (%) | 2/19 (11) | 2/8 (25) | 0/11 |
| **Laboratory Values (median) [IQR]** |  |  |  |
| WBC (median) [IQR] [4.0-11.0 E3/u] | (8.9) [6.2 - 10.7] | (5.75) [4.5 - 7.9] | (9.9) [7.7 - 12.1] |
| <4 | 1/50 (2) | 0/16 | 1/34 (3) |
| > 4 & < 11 | 29/50 (78) | 16/16 (100) | 23/34 (68) |
| >11 | 10/50 (20) | 0/16 | 10/34 (29) |
| Neutrophils (median) [IQR] [ Normal Range1.8-7 10E3/uL] | (6.5) [4.8-9.1] | (4.8) [3.25 - 6.05] | (8.4) [3.25 - 6.05] |
| > 1.8 & < 7 | 25/46 (54) | 14/16 (88) | 11/34 (37) |
| > 7 | 21/46 (46) | 2/16 (12) | 19/34 (63) |
| Lymphocytes (median) [IQR] [Normal range 1-3.4 10E3/uL] | (.95) [.7 - 1.3] | (1.2) [1 - 1.3] | (.85) [.6 - 1.3] |
| < 1 | 22/46 (48) | 3/16 (19) | 19/34 (63) |
| >1 < 3.4 | 24/46 (52) | 13/16 (81) | 11/34 (37) |
| Monocytes (median) [IQR] [0.2-0.8 10E3/uL] | (.50) [.7 - 1.3] | (0.5) [.35 - .65] | (.5) [.3 - .9] |
| Hemoglobin [12.0-16.0 g/dl] | (13.5) [11.3 - 15.1] | (15) [13.7 - 15.75] | (12.45) [11.1 - 14.2] |
| Platelets [ Normal range 150 - 400 E3/UL] | (234) [170 - 297] | 196 (152.5 - 293.5) | 236.5 (173 - 302) |
| INR [ Normal Range 0.8-1.3 ratio] | (1.18) [1.1 - 1.27] | (1.075) [1.02 - 1.18] | (1.19) [1.12 - 1.29] |
| D-Dimer [ Normal Range 0-500 ng/ml FEU] | (1252) [686 - 5865] | (677) [488.5 - 797] | (1882) [1047 - 7650] |
| Ferritin [ Normal Range 18-340 ng/ml] | (931.5) [441 - 1386] | (653) [26 - 929] | (1348) [476 - 2325] |
| CRP [ Normal Range <0.3 MG/DL] | (14.8) [7 - 18.5] | (8.85) [5.1 - 13.1] | (16.15) [8.5 - 22.25] |
| LDH [ Normal Range 117-224 unit/l] | (449) [324 - 590] | (236) [221 - 303] | (489) [377 - 619] |
| Lactate [ Normal Range 0.4-2.0 mmol/l] | (1.5) [1.3 - 2.2] | (1.35) [1 - 2.2] | (1.6) [1.3 - 2.2] |
| Procalcitonin [Normal Range <0.10 ng/ml] | (.26) [.11 - 1.58] | (.11) [.06 - .17] | (.95) [.16 - 3.07] |
| Sodium [ Normal Range 134 -144 mmol/L] | (136) [133-138] | (135.5) [132.5 - 137] | (136) [134 - 139] |
| Potassium [ Normal Range 3.5- 5.1 mmol/L] | (3.75) [3.5 - 4.1] | (3.55) [3.35 - 3.75] | (3.9) [3.6 - 4.1] |
| Chloride [ Normal Range 98-11 mmol/L] | (104) [101 - 107] | (104) [101.5 - 106] | (104) [100 - 108] |
| Creatinine [ Normal Range 0.50 - 1.40 mg/dl] | (.94) [.69 - 1.5] | (.715) [.625 - .94] | (1.1) [.76 - 4.03] |
| ALT [ Normal Range 6-68 Unit/l] | (36.5) [23-61] | (34.5) [22 - 61] | (37) [24 - 62] |
| AST [ Normal Range 14-67 Unit/l] | (45.5) [31 - 77] | (37) [26 - 74] | (57) [35.5 - 95] |
| ALP [ Normal Range 38-150 Unit/L] | (80) [68 - 133] | (78) [62 - 123] | (82) [68.5 - 135] |
| A1C [ Normal Range 4.4-5.6] | (7.3) [6.1 -10.4] | no observations | (7.3) [6.1 - 10.4] |
| Respiratory viruses other than COVID |  |  |  |
| RSV / Influenza A / Influenza B | 1/5 (20) | 1/4 (25) | 0/1 |
| Other Virus in Respiratory Panel | 5/30 (17) | 2/11 (18) | 3/19 (16) |
| Abnormal Chest x ray | 45/50 (90) | 13/16 (81) | 32/34 (94) |
| Median LOS in days | 7 days | 4 days | 13 days |

Legends: White blood cell ( WBC), International normalized ratio , C reactive protein ( CRP), Lactate dehydrogenase (LDH), Sodium , Aspartate aminotransferase (AST), Alanine transaminase (ALT), Alkaline phosphatase (ALP) , Hemoglobin A1c ( A1C), respiratory syncytial virus (RSV)

Median WBC counts were 8.9(6.2-10.7). On differential analysis 46% (25/46) had elevated neutrophil counts, and 48% (22/46) had low lymphocytes counts. Median D dimer, Ferritin, CRP, LDH were all elevated at presentation but lactic acid and procalcitonin were within normal limits. 43.4% (20/46) patients has elevated AST, 21.7% (10/46) has elevated ALT, 10.8 % (5/46) had elevated ALP and 13.0% (6/46) had elevated total bilirubin.

All patients tested for COVID-19 and out of them who had Respiratory pathogen panels tested 6/31(19.3%) tested positive for other respiratory pathogens; 3 were positive for adenovirus, 1 for metapneumovirus, 1 for influenza and 1 for rhinovirus. 1 out of 6 patients with coinfection with another respiratory virus died.

97.5% (40/41) patients had negative blood cultures and only 2 % (1/41) had a positive blood culture and received antibiotics. Sputum cultures were negative in 92.3% (12/13) patients. Only 1/38(2%) have a positive nasal MRSA swab and none of the patient tested for legionella (3) and streptococcus pneumonia antigen (8) in urine tested positive. 10% (5/50) had a negative initial chest x-ray.

### Management

37/49(75.5%) patient received azithromycin, 38/48(70.8%) received ceftriaxone and 19/49(38.8%) received Hydroxychloroquine. 1 patient received Remdesivir and 1 patient received Tocilizumab. 7/50(14%) received steroids. Of these 7,1 was on chronic steroids which was continued, 1 was given steroids for 1 day for suspected asthma exacerbation and remaining 5 patients received it for refractory septic shock. Out of 6 patients in the ICU who got stress dose steroids 1 was still admitted at the end of the study and 5 of them died (100%) though these patients were much sicker to start with. 20/34(58.8%) patients going to ICU required vasopressor support. Median number of days they were on vasopressor was 6(range 2-18days)

Of admitted patients, 17/50(34%) were directly admitted to ICU and 17/50(34%) were first admitted to floor and then transferred to the ICU. Of these ICU patients 28/34(82.4%) required invasive mechanical ventilation and 5/34(14.7%) were managed with high flow nasal cannula. Only one patient was transferred to ICU for continuous renal replacement therapy (CRRT). Patients spent a median of 2 days on the floor prior to ICU transfer, median length of stay in the ICU was 7 days. On comparing characteristics of patients, patients with diabetes, and higher lactate dehydrogenase on admission were more likely to require ICU level of care. (Table III)

**Table III.** Comparing Floor vs. ICU Patients (Chi-Squared)

| Table III Comparing Floor vs. ICU Patients (Chi-Squared) |  |  |  |
| --- | --- | --- | --- |
| Clinical characteristics | Floor patients | ICU patients | P value |
| Age | (n/N) |  |  |
| 20 -40 years | 3/16 (19%) | 10/34 (29) | 0.724 |
| 41 - 64 years | 8/16 (50) | 15/34 (44) |  |
| > 65 years | 5/16 (31) | 9/34 (28) |  |
| Sex |  |  |  |
| Male | 8/16 (50) | 15/34 (44) | 0.697 |
| Female | 8/16 (50) | 19/34 (56) |  |
| HTN | 3/16 (19%) | 14/34 (41) | 0.118 |
| Diabetes | 2/16 (13) | 16/34 (47) | 0.018 |
| Asthma | 1/16 (6) | 5/34 (15) | 0.391 |
| Alcoholic Cirrhosis | 0/16 | 2/34 (6) | 0.322 |
| Alcohol use (active) | 0/16 | 6/34 (18) | 0.073 |
| Obesity | 5/12 (42) | 15/27 (56) | 0.423 |
| Symptom Onset |  |  |  |
| 1-5 days | 3/15 (20) | 14/26 | 0.02 |
| 6-10 days | 10/15 (66) | 5/26 (19) |  |
| 11-15 days | 2/15 (13) | 5/26 (19) |  |
| >15 days | 0/15 | 2/26 (8) |  |
| Lab Values |  |  |  |
| D-Dimer ng/mL |  |  |  |
| Normal ( 0- 500) | 2/8 (25) | 1/25 (4) | 0.07 |
| Abnormal* | 6/8 (75) | 24/25 (96) |  |
| Ferritin ng/mL |  |  |  |
| Normal (30 - 530) | 2/7 (29) | 5/15 (33) | 0.823 |
| Abnormal * | 5/7 (71) | 10/15 (66) |  |
| CRP mg/dL |  |  |  |
| Normal (<0.3) | 0/10 | 0/10 | ~ |
| Abnormal* | 10/10 (100) | 28/28 (100) |  |
| ESR mm/hr |  |  |  |
| Normal ( 0-33) | 3/8 (38) | 2/22 (9) | 0.06 |
| Abnormal * | 5/8 (62) | 20/22 (91) |  |
| LDH unit/L |  |  |  |
| Normal (117-224) | 2/6 (33) | 0/23 (0) | 0.004 |
| Abnormal* | 4/6 (66) | 23/23 (100) |  |
| Creatinine |  |  |  |
| Normal (0.5-1.4) | 15/16 (94) | 20/34 (59) | 0.012 |
| Abnormal* | 1/16 (6) | 14/34 (41) |  |

\* abnormal category defined as lab values outside reference ranges
C reactive protein ( CRP), Lactate dehydrogenase (LDH), Erythrocyte sedimentation rate (ESR), Hypertension (HTN)

## Outcomes

### Mortality

No patient deaths were reported on the floor. Of 34 patients in the ICU 13 died while 6 are still receiving care in the hospital, with an overall mortality of 30.2% (13/43). Out of 13 patients who died, 2 were on hemodialysis (HD), 11/13(84%) patients had acute kidney injury [AKI] and required CRRT or HD. (Graph I)

**Graph I.**
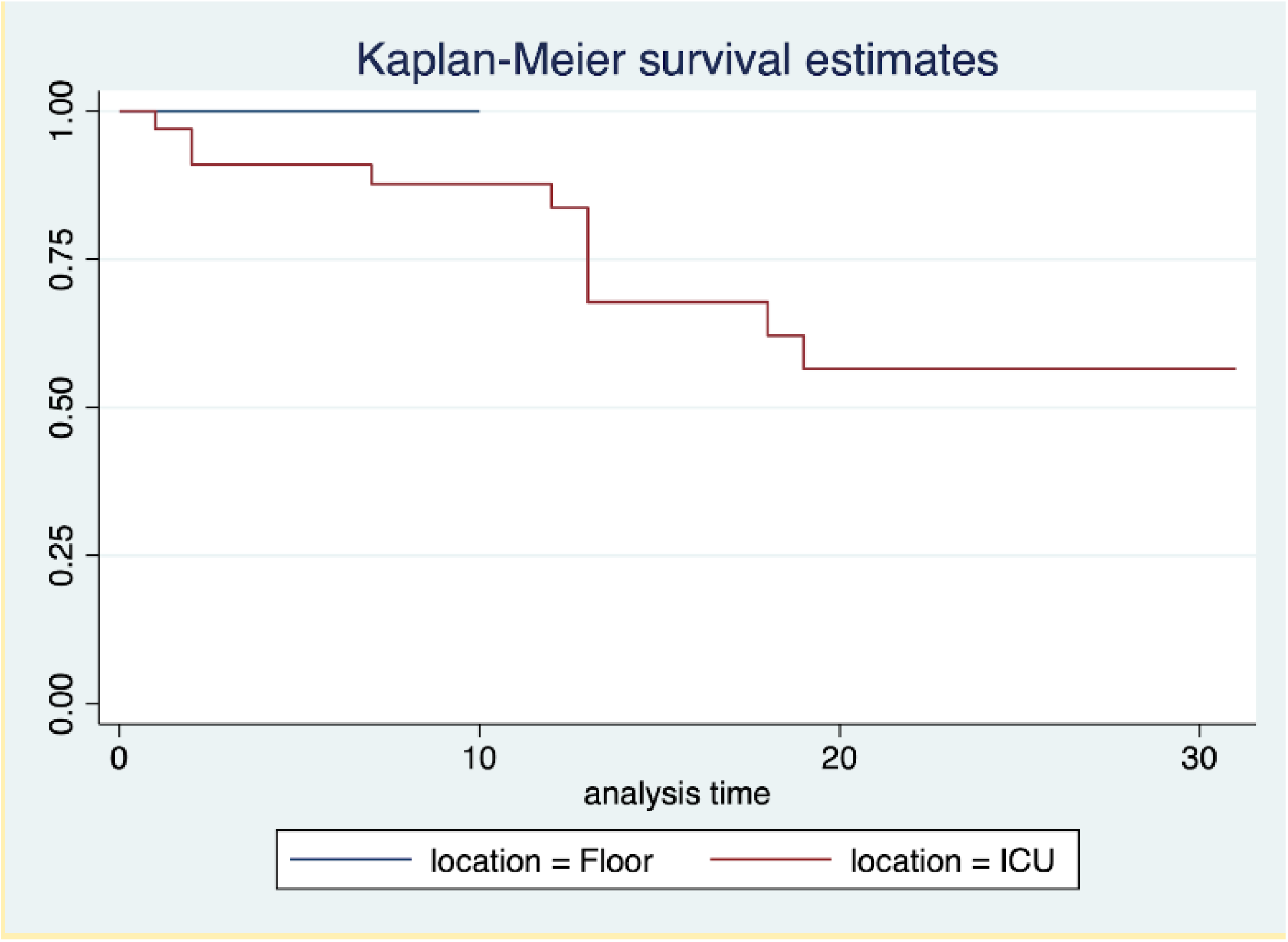
Kaplan-Meier survival estimates

### Complication

9/50(18%) patients had no complications. 1/50(2%) developed a pulmonary embolism, 21/50 (42%) deve006Coped septic shock and 20/50 (40%) required vasopressor support. 4/50(8%) experienced cardiopulmonary arrest with, 1 STEMI (2%) and 4(8%) Non-STEMI. 11/50(22%) developed AKI. 2/50(4%) experienced cerebrovascular events and 3/50(6%) developed intracranial bleeds (they all were on DVT prophylaxis with heparin), 11/50(22%) were encephalopathic (mostly metabolic encephalopathy) and 2/50(4%) experienced anoxic brain injury. 4/50(8%) developed acute liver injury and 1 patient had gastrointestinal bleeding. In the recovery phase, 4/50(8%) patients reported dysphagia and 7/50(14%) patients suffered from severe deconditioning.

### Length of stay and discharge disposition

The median length of stay is 7 days (Range 1-31days), for floor patients 4 days and ICU patients 13 days. Out of 43 patients who completed their clinical course 24/43(58.1%) were discharged home, 5/43(11.6%) went to rehabilitation facilities and 30.2% died. 16/30(53.3%) required oxygen on discharge. (Graph II)

**Graph II.**
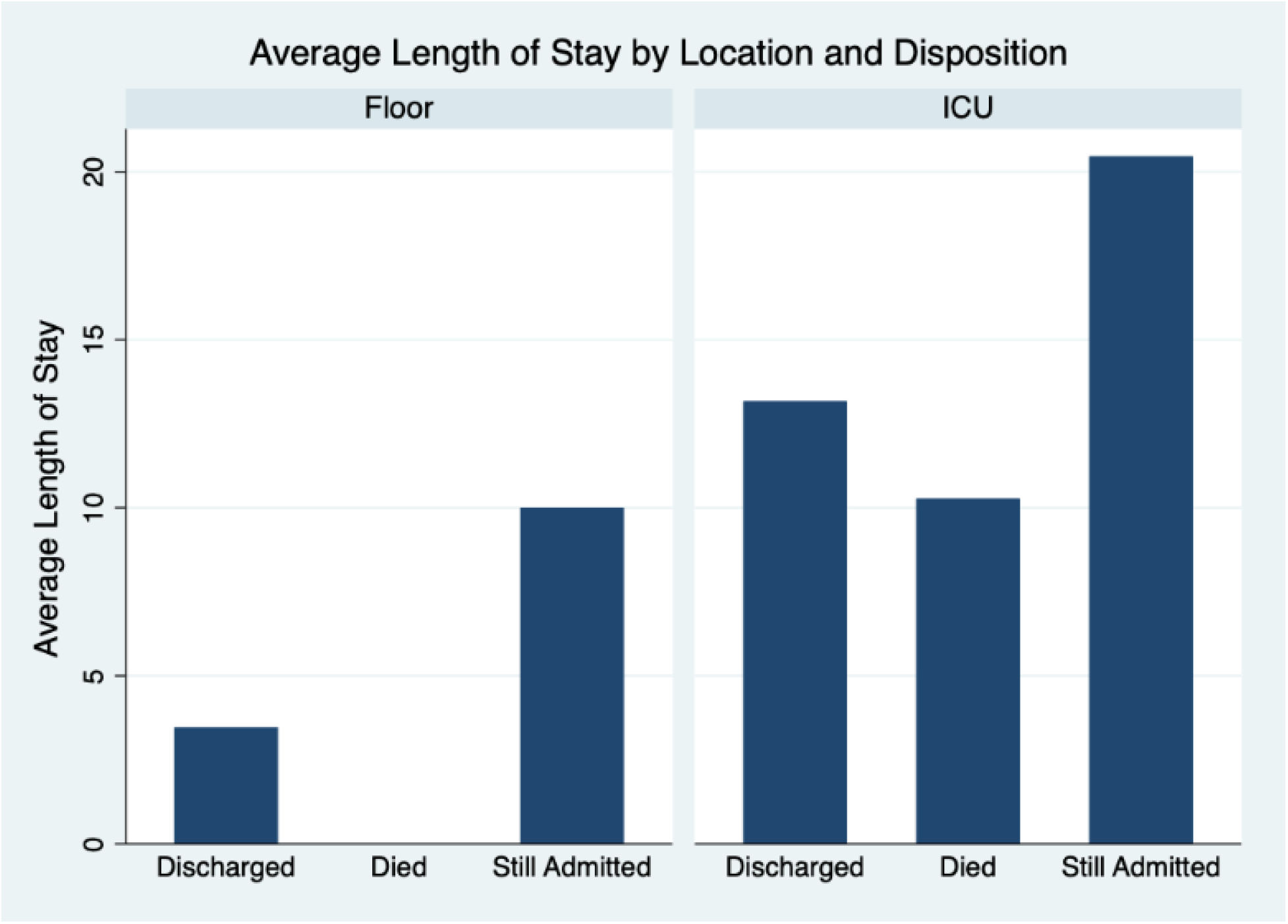
Length of stay, Floor v/s ICU

## Discussion

We present our early experience in managing 50 COVID-19 patients at a tertiary care Academic Hospital in the South-western United States. We included patients both from the general medical floor and ICU and extensive chart review was performed. While our proportion of female patients was more than that reported elsewhere[^3,4,5^], it is in accordance with the demographics of New Mexico[^6^]

The most common presenting symptoms in our patient's population was subjective fever, cough which was dry in nature and shortness of breath. Only one fourth of the patient population was actually febrile during hospitalization. This is different from larger case series published from China^7^ but more in line with US case series[^5 4^]. Also, 90% of the patients had X-Ray findings described mainly as bilateral or basilar infiltrates. Based on this, suggest that a history of subjective fever instead of documented temperatures, along with cough, shortness of breath and abnormal x-ray can be used as an initial clinical tool to identify and triage patients, at least till a faster and more accessible modality to screen for coronavirus is made available. Co-infection with other respiratory viruses should not preclude COVID-19 testing.

The mean duration of symptoms onset to presentation to hospital for admission was over a week and hospitalization occurred even though more than half of the patients sought initial outpatient healthcare earlier. Similar to previous reports, our patients had several comorbidities such as obesity, diabetes, hypertension and smoking, and required oxygen at admission

Most patients received 2 or more antibiotics targeted towards community acquired pneumonia however only 1/50 patient had a positive blood culture. Patients also received azithromycin and hydroxychloroquine as part of an ongoing clinical trial[^8^]. Our study was not powered enough to give outcomes but larger studies are needed to see if antibiotics for community acquired pneumonia coverage will make any difference in outcomes.

Diabetics were more likely to be admitted to the ICU, as well as patients with high baseline LDH. Over 80 percent of patients who died developed AKI and required CRRT. Thus, development of AKI is a bad prognostic sign in patients with COVID infection as cited by other studies.

Our study has several notable limitations. First some cases had incomplete documentation of clinical symptoms, missing laboratory testing, or both. Second, our sample size is small and being a tertiary care teaching referral hospital, our patient population was sick to start with so mortality data may not be representative of the general population. Third, this data is representative of the early phase of SARS-cov-2 transmission in New Mexico. Several factors will change over time which might affect the outcome in future and hopefully a better one.

## Data Availability

all data was extracted from electronic medical record and stored in red cap in de identified form

## Abbreviation

COVID-19 - coronavirus disease 2019, UNM - University of New Mexico, OSH - Outside hospital, SNF - Skilled nursing facility, Ra - Room air, Sats - oxygen saturation, ICU - Intensive care Unit, WBC -white blood cell counts, CRP- C reactive protein, LDH - Lactate dehydrogenase, AST-Aspartate aminotransferase, ALT- Alanine transaminase, ALP - Alkaline phosphatase, MRSA- Methicillin-resistant Staphylococcus aureus, HD - Hemodialysis, CRRT -Continuous Renal Replacement Therapy, AKI - Acute kidney injury, STEMI - ST-elevation myocardial infarction, Non-STEMI - Non-ST-elevation myocardial infarction, HTN -Hypertension, ESR- Erythrocyte sedimentation rate, WBC - white blood cell counts, MRSA - methicillin resistant staph aureus, AKI - acute Kidney injury, STEMI - St Segment elevation myocardia infraction, Non STEMI - Non St Segment elevation myocardia infraction, DVT - Deep venous thrombosis.

